# Impact of the COVID-19 pandemic on children and adolescents: determinants and association with quality of life and mental health – A cross-sectional study

**DOI:** 10.1101/2022.11.18.22282491

**Authors:** Viviane Richard, Roxane Dumont, Elsa Lorthe, Andrea Loizeau, Hélène Baysson, María-Eugenia Zaballa, Francesco Pennacchio, Rémy P. Barbe, Klara M. Posfay-Barbe, Idris Guessous, Silvia Stringhini, SEROCoV-KIDS study group

**Author notes:** CORRESPONDENCE TO Silvia Stringhini, Unité d’épidémiologie populationnelle, Rue Jean-Violette 29, 1205 Genève, Switzerland. Authors contributed equally to this work (Idris Guessous, Silvia Stringhini). SEROCoV-KIDS study group Deborah Amrein, Isabelle Arm-Vernez, Andrew S Azman, Antoine Bal, Michael Balavoine, Rémy P Barbe, Hélène Baysson, Julie Berthelot, Patrick Bleich, Livia Boehm, Gaëlle Bryand, Viola Bucolli, Prune Collombet, Alain Cudet, Vladimir Davidovic, Carlos de Mestral Vargas, Paola D'Ippolito, Richard Dubos, Roxane Dumont, Isabella Eckerle, Nacira El Merjani, Marion Favier, Natalie Francioli, Clément Graindorge, Idris Guessous, Séverine Harnal, Samia Hurst, Laurent Kaiser, Omar Kherad, Julien Lamour, Pierre Lescuyer, Arnaud G. L’Huillier, Andrea Jutta Loizeau, Elsa Lorthe, Chantal Martinez, Stéphanie Mermet, Mayssam Nehme, Natacha Noël, Francesco Pennacchio, Javier Perez-Saez, Anne Perrin, Didier Pittet, Klara M Posfay-Barbe, Jane Portier, Géraldine Poulain, Caroline Pugin, Nick Pullen, Viviane Richard, Frederic Rinaldi, Deborah Rochat, Cyril Sahyoun, Irine Sakvarelidze, Khadija Samir, Hugo Alejandro Santa Ramirez, Jessica Rizzo, Stephanie Schrempft, Claire Semaani, Silvia Stringhini, Stéphanie Testini, Yvain Tisserand, Deborah Urrutia Rivas, Charlotte Verolet, Jennifer Villers, Guillemette Violot, Nicolas Vuilleumier, Sabine Yerly, María-Eugenia Zaballa, Christina Zavlanou.

## Abstract

**Background:** The medium-term impact of the COVID-19 pandemic on the wellbeing of children and adolescents remains unclear. More than two years into the pandemic, we aimed to quantify the frequency and determinants of having been severely impacted by the COVID-19 pandemic and estimate its impact on health-related quality of life (HRQoL) and mental health.

**Methods:** Data was drawn from a population-based cohort of children and adolescents, recruited between December 2021 and June 2022, in Geneva, Switzerland. We measured the impact of the pandemic via the Coronavirus impact scale, which assesses the multidimensional impact of the pandemic at the child and family level through parent’s report. A score higher than one standard deviation above the mean was deemed a severe impact. Parents additionally reported about their offspring HRQoL and mental health with validated scales. Determinants of having been severely impacted were assessed with logistic models, as were the associations between having experienced a severe impact and poor HRQoL or mental health.

**Results:** Out of 2101 participants aged 2-17, 12.7% had experienced a severe pandemic impact. Having a lasting health condition, a pandemic-related worsening of lifestyle habits or an unfavorable family environment were associated with having been severely impacted by the pandemic. Participants who had experienced a severe pandemic impact were more likely to present poor HRQoL (aOR=3.1; 95%CI: 2.3-4.4) and poor mental health (aOR=3.9; 95%CI: 2.5-6.2).

**Conclusions:** The COVID-19 pandemic may have persistent consequences on the wellbeing of children and adolescents, especially among those with health and family vulnerabilities.

## INTRODUCTION

As a consequence of measures enforced to control the spread of the COVID-19 pandemic, children and adolescents faced various life disruptions, including online or interrupted education (1), delayed healthcare seeking and provision (2), decrease in contact with peers and relatives outside the household, or routine modification with prolonged screen time and reduced organized physical activity (3).

If the pandemic has affected most, if not all, of the world population, its impact is likely to have been heterogeneous. Early evidence showed that children and adolescents with an existing health condition seemed particularly affected (4,5), possibly because they were more exposed to disruptions in routine healthcare or to an increase in their caregiver’s stress due to the lack of specialized care provision (6). Social connectedness and family support also appeared key to mitigating the impact of the pandemic (7). However, the pandemic strongly modified some families’ circumstances, since some caregivers lost their income, while others experienced an increase in workload or the introduction of remote working together with a disruption of childcare services (8). It may have been challenging for strained caregivers to navigate the changing pandemic context while having the emotional availability to nurture harmonious family relationships and provide adequate support to their children (8). Finally, a confirmed SARS-CoV-2 infection, although usually associated with mild COVID-19 among children, may have represented a disturbing and stressful experience because of potential isolation, stigma or school disruption (9)

Routine changes and stressful circumstances caused by the pandemic are thought to have affected children’s and adolescents’ well-being. Systematic reviews conducted during the first year of the pandemic found a decrease in their health-related quality of life (HRQoL) and mental health, with a worsening trend over time (10,11). More recent studies showed a stabilization or even a slight improvement in their well-being two years into the pandemic (12,13); it could be linked to both the development of strategies to cope with the pandemic situation and the lifting of sanitary measures. On the other hand, the burden may remain high for those who experienced multiple pandemic stressors (14). However, the prevalence and determinants of persistent effects of the pandemic on children’s and adolescents’ HRQoL and mental health remain largely unknown, especially in countries with relatively light restrictions, such as Switzerland (15). Indeed, schools remained open after closing during a 10-week semi-lockdown in spring 2020, while remote working, varying limitations in the size of gathering and quarantine after contact with a COVID-19 case were maintained. Inland travel was never limited; non-essential shops, sport and cultural places intermittently closed according to the epidemiological situation. All measures were lifted on February 3^rd^ 2022.

In this study, we aimed to 1) quantify the frequency of having been severely impacted by the COVID-19 pandemic among children and adolescents (hereafter referred to as children), 2) identify related determinants, and 3) evaluate the association of a severe impact with HRQoL and mental health two years into the pandemic.

## METHODS

### Study design

Data was drawn from the baseline assessment of the SEROCoV-KIDS study, a population-based prospective cohort study, which aims at evaluating the impacts of the COVID-19 pandemic on the health and well-being of children.

Eligibility criteria were (1) having been (or having a sibling) selected from random samples of the Geneva population provided by the Swiss Federal Office of Statistics or the Cantonal Population and Migration Office, or being part of a randomly selected family participating in a previous seroprevalence study (16), (2) being between 6 months and 17 years of age and, (3) living in the canton of Geneva at the time of enrollment. Participation rate varied according to previous participation (returning participants: 42.1%; newly selected ones: 9.1%). Among new participants, participation rate was higher among older children and adolescents (10.5% for 6-13 years; 9.5% for 14-17 years) compared with children aged 1-5 years (7.1%).

Baseline assessment took place between December 2021 and June 2022. All participants were invited to perform a SARS-CoV-2 serological test from blood drawing. Further information was collected through an online and secured platform. One parent or referent adult per family filled out a socio-demographic, health, and lifestyle questionnaire for each participating child, and one for themselves. Adolescents (aged 14 or older) completed a questionnaire about their lifestyle.

### Study population

For the current analyses, we selected children with complete information on the main variable of interest (impact of the COVID-19 pandemic) and born before January 1^st^ 2020, i.e. at least two months before the start of the pandemic in Switzerland (Additional file, figure 1).

**Fig 1.**
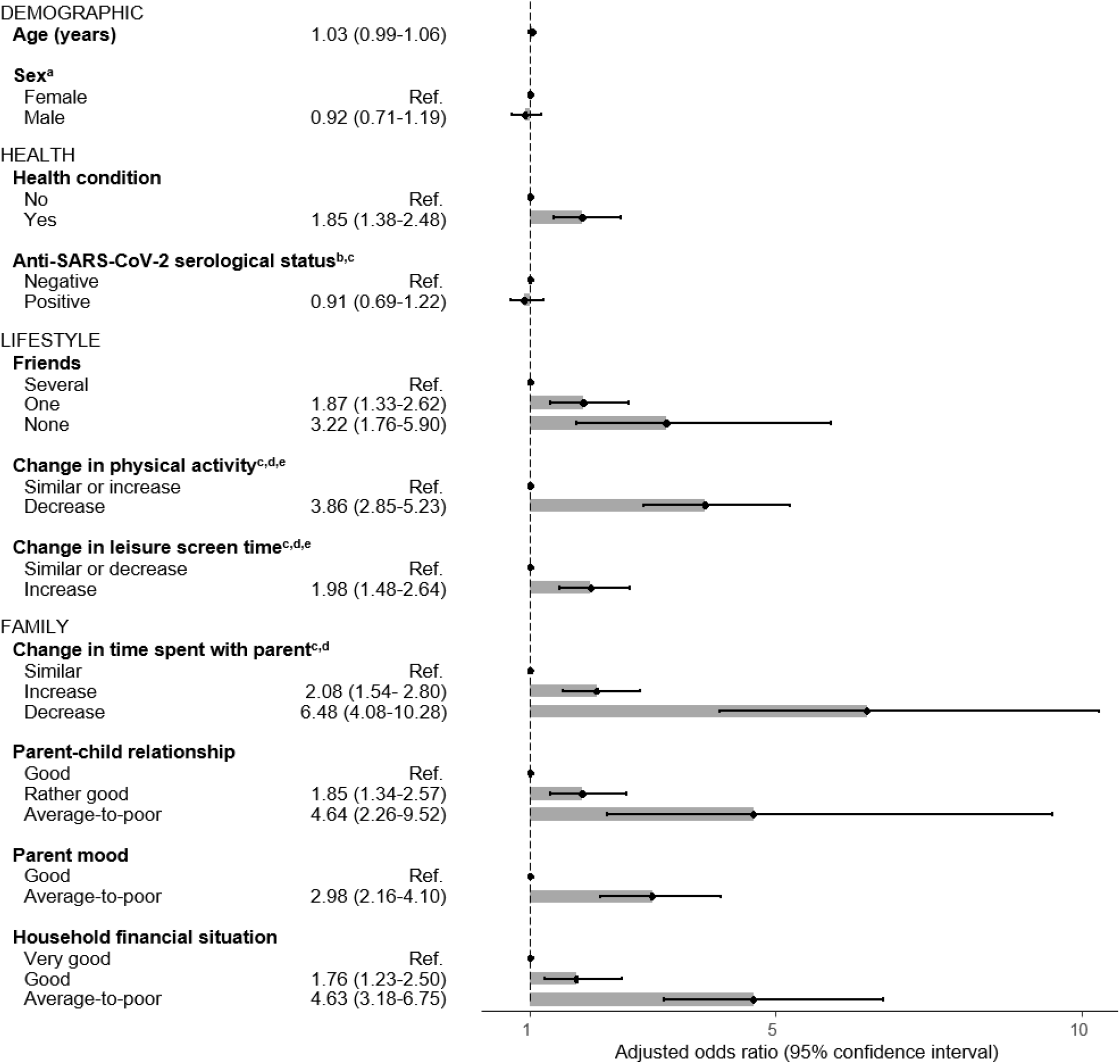
Determinants of having been severely impacted by the COVID-19 pandemic among children and adolescents (n=2043). Results from generalized estimating equations taking the household clustering into account and adjusted for age and sex. Model for parent mood adjusted for age, sex, health condition and financial situation; model for parent-child relationship adjusted for age, sex and parent mood. ^a^ Sex category “Other” not included because of too small number of observations. ^b^ Regression based on participants with available anti-SARS-CoV-2 serology (n=1966). ^c^ Category Unknown/Not applicable, Unknown/No answer or Undetermined not shown. ^d^ Participant-reported change as a result of the COVID-19 pandemic. ^e^ Similar level and increase in physical activity as well as similar level and decrease in screen time are grouped. When analyzed separately, coefficients were of comparable magnitude

### Measures

#### COVID-19 impact

The Coronavirus impact scale was developed to evaluate the multidimensional impact of the COVID-19 pandemic on adults (17). We adapted the scale to measure the impact of the pandemic on children through parental report. Parents rated how the pandemic impacted their offspring or family on a 4-point Likert scale (no change, mild, moderate, severe) in the following domains: routine, income, food access, physical and mental healthcare access, access to social and family support, stress, and family discord. The 8 items were summed up to compute an overall impact score, a higher value indicating a more severe impact; the internal consistency was adequate (α=0.63), similar to the original scale (17). The measure was dichotomized and a score higher than one standard deviation above the study population mean defined as a severe pandemic impact. Sensitivity analyses were performed, with a score lying within the third tertile of the study population considered as severe.

#### Health-related quality of life

HRQoL was assessed with the French version of the PedsQL Short Form (18). HRQoL was self-reported by adolescents aged 14 and above and parent-reported for children aged between 2 and 13. The 15 items of the scale were added up and linearly transformed on a scale between 0 and 100, a higher score indicating a better HRQoL. Internal consistency was good (α=0.85). Physical and psychosocial functioning were further assessed using the corresponding subscales (α=0.93 and α=0.81, respectively). Poor HRQoL, physical or psychosocial functioning were defined using thresholds compatible with a severe health condition (19).

#### Mental health

Mental health was measured with the French version of the parent-reported SDQ (20). Items were summed up following scoring instructions. The overall score ranged from 0 to 40, with an adequate internal consistency (α=0.78); a poor mental health was defined using clinical threshold defined as the 90^th^ percentile of the score distribution based on United Kingdom normative data (https://sdqinfo.org). As recommended for community samples (21), we computed the internalizing score by adding the emotional and peer problems subscales (α=0.68), and the externalizing score by combining the conduct problems and hyperactivity subscales (α=0.78). The internalizing and externalizing scores were dichotomized with cut-offs defined as the addition of the clinical cut-off of the corresponding subscales.

#### Socio-demographic, health, behavioral and family measures

Age and sex of children were collected, as well as health indicators such as the presence of a professionally diagnosed physical health condition or mental behavioral and neurodevelopmental disorder lasting over 6 months (referred to hereafter as health condition, Additional file, table 1), and a previous SARS-CoV-2 infection. Since vaccines available in Switzerland do not trigger an immune response against the SARS-CoV-2 nucleocapsid (N) protein, a serological test detecting anti-SARS-CoV-2 N antibodies (Elecsys; Roche Diagnostics, Rotkreuz, Switzerland) was used to objectively assess a previous SARS-CoV-2 infection, with a cut-off index ≥ 1.0 considered as positive, as indicated by the manufacturer. Lifestyle changes as a result of the pandemic were also collected; change in leisure screen time was parent-reported for children aged between 1 year at pandemic onset and 13 years, and self-reported by adolescents aged from 14 years, while change in physical activity time was parent-reported for children and adolescents aged at least 2 years at the pandemic onset. Parents additionally reported about their offspring’s number of close friends if aged 6 years or older.

**Table 1.** Characteristics of children and adolescents according to the pandemic impact

|  | Total<br>(n=2101) | COVID-19 pandemic impact |  | P-value |
| --- | --- | --- | --- | --- |
|  |  | None or mild<br>(n=1834) | Severe<br>(n=267) |  |
| **Age (median [IQR])** | 10 (7-13) | 10 (7-13) | 10 (7-14) | 1.000 |
| **Sex** |  |  |  |  |
| Female | 1035 (49.3) | 901 (49.1) | 134 (50.2) | 1.000 |
| Male | 1063 (50.6) | 930 (50.7) | 133 (49.8) |  |
| Other | 3 (0.1) | 3 (0.2) | 0 (0.0) |  |
| **Physical or mental health condition** |  |  |  |  |
| No | 1653 (78.7) | 1473 (80.3) | 180 (67.4) | <0.001 |
| Yes | 447 (21.3) | 360 (19.6) | 87 (32.6) |  |
| Missing | 1 (0.0) | 1 (0.1) | 0 (0.0) |  |
| **Anti-SARS-CoV-2 N serological status** |  |  |  |  |
| Negative | 611 (29.1) | 530 (28.9) | 81 (30.3) | 1.000 |
| Positive | 1387 (66.0) | 1219 (66.5) | 168 (62.9) |  |
| Undetermined | 18 (0.9) | 17 (0.9) | 1 (0.4) |  |
| Missing | 85 (4.0) | 68 (3.7) | 17 (6.4) |  |
| **Number of friends** |  |  |  |  |
| Several | 1364 (64.9) | 1215 (66.2) | 149 (55.8) | <0.001 |
| One | 305 (14.5) | 249 (13.6) | 56 (21.0) |  |
| None | 58 (2.8) | 41 (2.2) | 17 (6.4) |  |
| Unknown / Not applicable <sup>a</sup> | 374 (17.8) | 329 (17.9) | 45 (16.9) |  |
| **Change in physical activity time <sup>b</sup>** |  |  |  |  |
| Similar | 1333 (63.4) | 1227 (66.9) | 106 (39.7) | <0.001 |
| Increase | 170 (8.1) | 141 (7.7) | 29 (10.9) |  |
| Decrease | 341 (16.2) | 244 (13.3) | 97 (36.3) |  |
| Unknown / Not applicable <sup>c</sup> | 257 (12.2) | 222 (12.1) | 35 (13.1) |  |
| **Change in leisure screen time <sup>b</sup>** |  |  |  |  |
| Similar | 886 (42.2) | 806 (43.9) | 80 (30.0) | <0.001 |
| Increase | 975 (46.4) | 813 (44.3) | 162 (60.7) |  |
| Decrease | 23 (1.1) | 21 (1.1) | 2 (0.7) |  |
| Unknown / Not applicable <sup>c</sup> | 167 (7.9) | 154 (8.4) | 13 (4.9) |  |
| Missing | 50 (2.4) | 40 (2.2) | 10 (3.7) |  |
| **Change in time spent with parent <sup>b</sup>** |  |  |  |  |
| Similar | 1309 (62.3) | 1199 (65.4) | 110 (41.2) | <0.001 |
| Increase | 640 (30.5) | 539 (29.4) | 101 (37.8) |  |
| Decrease | 97 (4.6) | 61 (3.3) | 36 (13.5) |  |
| Unknown | 55 (2.6) | 35 (1.9) | 20 (7.5) |  |
| **Parent-child relationship** |  |  |  |  |
| Good | 1723 (82.0) | 1541 (84.0) | 182 (68.2) | <0.001 |
| Rather good | 338 (16.1) | 268 (14.6) | 70 (26.2) |  |
| Average-to-poor | 40 (1.9) | 25 (1.4) | 15 (5.6) |  |
| **Parent mood** |  |  |  |  |
| Good | 1832 (87.2) | 1644 (89.7) | 186 (69.7) | <0.001 |
| Average-to-poor | 267 (12.7) | 187 (10.2) | 80 (30.0) |  |
| Missing | 2 (0.1) | 1 (0.1) | 1 (0.4) |  |
| **Household financial situation** |  |  |  |  |
| Very good | 732 (34.8) | 680 (37.1) | 52 (19.5) | <0.001 |
| Good | 864 (41.1) | 759 (41.4) | 105 (39.3) |  |
| Average-to-poor | 360 (17.1) | 264 (14.4) | 96 (36.0) |  |
| Unknown / No answer | 142 (6.8) | 129 (7.0) | 13 (4.9) |  |
| Missing | 3 (0.1) | 2 (0.1) | 1 (0.4) |  |
| **Health-related quality of life** |  |  |  |  |
| Average to good | 1843 (87.7) | 1650 (90.0) | 193 (72.3) | <0.001 |
| Poor | 208 (9.9) | 144 (7.9) | 64 (24.0) |  |
| Missing | 50 (2.4) | 40 (2.2) | 10 (3.7) |  |
| **Mental health** |  |  |  |  |
| Average to good | 1994 (94.9) | 1764 (96.2) | 230 (86.1) | <0.001 |
| Poor | 105 (5.0) | 68 (3.7) | 37 (13.9) |  |
| Missing | 2 (0.1) | 2 (0.1) | 0 (0.0) |  |

Results are numbers (%) for categorical variables and median (interquartile range) for continuous ones. P-values are from Chi-squared test for categorical variables and Kruskal-Wallis test for continuous ones, adjusted for multiple comparison with the Bonferroni method.
<sup>a</sup> Not applicable if younger than 6 years. <sup>b</sup> Reported change as a result of the pandemic. <sup>c</sup> Not applicable if stated so by parent, or if younger than 1 year for leisure screen time change or 2 years for physical activity at pandemic onset

Family-level information was also collected. Parent-child relationship was reported by the parent with the question “In general, how would you evaluate your relationship with your child?”; possible answers were “good”, “rather good”, as well as “average”, “rather poor” and “poor”, which were grouped into an “average-to-poor” category. Effect of the pandemic on the time parent and child spend together was evaluated with the question “Did the pandemic have an impact on the time you currently spend with your child?”. Parental mood was defined as “good” if parents answered “very good” or “good” to the question “In general, how would you assess your mood?”, and “average-to-poor” for the answers “average”, “poor” and “very poor”. The household financial situation was considered as “very good” if parents reported a comfortable financial situation in which they could easily save money, “good” if they could cover their needs and face unexpected minor expenses, and “average-to-poor” if unexpected expenses could put them into financial difficulties or if they were not able to cover their expenses and needed external support.

### Statistical analyses

The prevalence of having been severely impacted by the pandemic and of poor HRQoL or mental health were standardized to the age and sex distribution of the 2021 Geneva population (22). Differences in the distribution of the pandemic impact according to children’s characteristics were tested with Chi-squared test for categorical variables and Kruskal-Wallis test for continuous variables, adjusted for multiple comparison with the Bonferroni method.

Each of the above-mentioned variables was separately assessed as a possible determinant for having been severely impacted by the pandemic using logistic regression models adjusted for age and sex. To account for potential confounders, specific adjustments were made. The model for parent mood was additionally adjusted for health condition of the child and household financial situation, and the model for parent-child relationship was further adjusted for parent mood (Additional file, figure 2). The association between having been severely impacted by the pandemic and HRQoL or mental health was estimated with logistic models first adjusted for age and sex, and subsequently for health, lifestyle and family factors as potential confounders. To take the household clustering of data into account, analyses were based on generalized estimating equations performed with the R geepack (23), following a binomial distribution.

**Fig 2.**
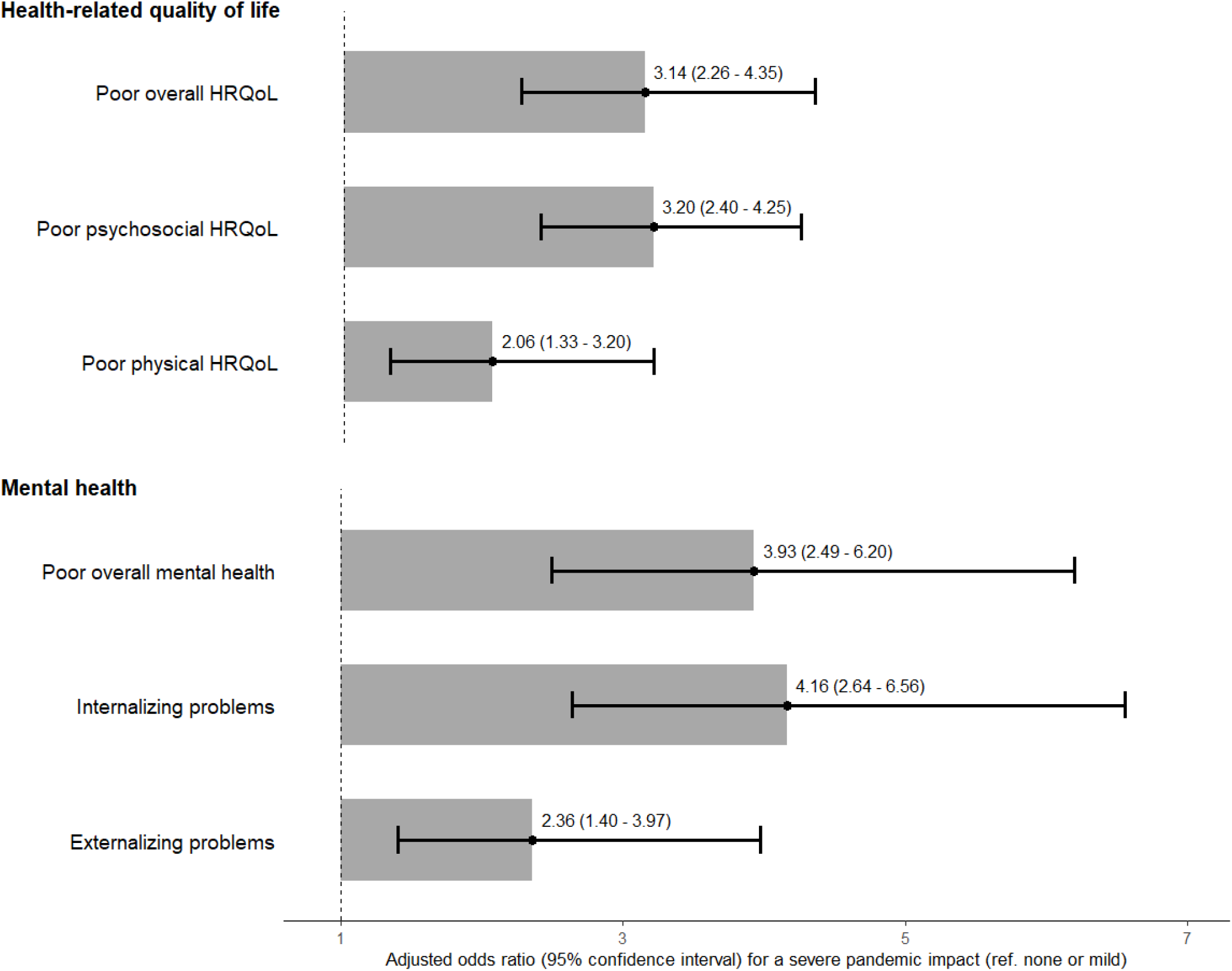
Association between having been severely impacted by the COVID-19 pandemic and health-related quality of life (HRQoL) or mental health of children and adolescents. Results from generalized estimating equations taking the household clustering into account, adjusted for sex, age and health condition. Reference levels are an average-to-good HRQoL and mental health, and no internalizing/externalizing problems (n=2043)

Overall, 132 (6.3%) participants had at least one missing data, that was deemed missing at random and imputed using multiple imputation by chained equations with the R mice package (24). Predictive mean matching was used for numerical variables, logistic regression for dichotomous ones and polytomous regression for categorical ones. Variables included in the above-described models, as well as children’s general health, mood, country of birth and daily physical activity time were added as predictors for the imputation.

Analyses were performed with R (version 4.1.2) and statistical significance was defined at a confidence level of 95%.

## RESULTS

### Descriptive analyses

The resulting sample comprised 2101 children and adolescents aged between 2 and 17 years (median: 10.0; IQR: 7-13), with 49.3% girls (Table 1). Routine, and social and family contact were frequently reported as having been disrupted by the pandemic, while an impact on food and physical or mental healthcare access was only reported for a minority (Additional file, figure 3). When standardized to the age and sex distribution of the Geneva population, 12.7% of participants had been severely impacted by the COVID-19 pandemic (mean score: 3.7), 11.5% had a poor HRQoL (mean score: 83.8), and 5.1% had a poor mental health (mean score: 7.2). Although HRQoL and mental health were slightly poorer among participants recruited after the easing of measures, the difference was not clinically relevant (Additional file, table 2).

#### Determinants of having been severely impacted by the COVID-19 pandemic

Determinants for having been severely impacted by the COVID-19 pandemic included having only one (aOR=1.9; 95%CI: 1.3-2.6) or no friends (aOR=3.2; 95%CI: 1.8-5.9), compared to having several, and suffering from a health condition (aOR=1.9; 95% CI: 1.4-2.5; Fig. 1). Lifestyle changes as a result of the pandemic, such as an increase in leisure screen time (aOR=2.0; 95%CI: 1.5-2.6) or a decrease in time spent engaging in physical activity (aOR=3.9; 95%CI: 2.9-5.2), were also associated with experiencing a severe impact. Finally, children living in an unfavorable family environment characterized by an average-to-poor parent-child relationship (aOR=4.6; 95%CI: 2.3-9.5), average-to-poor parent mood (aOR=3.0; 95%CI: 2.2-4.1) or average-to-poor financial situation (aOR=4.6; 95%CI: 3.2-6.8), were also more likely to have been severely impacted by the pandemic compared with those living in better circumstances. A change in time spent with the parent as a result of the pandemic, whether an increase (aOR=2.1; 95%CI: 1.5-2.8) or decrease (aOR=6.5; 95% CI: 4.1-10.3), was associated with having experienced a severe pandemic impact. The anti-SARS-CoV-2 N serological status, was not associated with the pandemic impact (P-value>0.05). Results with imputed data and with different definition of a severe impact were of similar magnitude (Additional file, table 3).

#### Association between having been severely impacted by the COVID-19 pandemic and health-related quality of life or mental health

Having been severely impacted by the COVID-19 pandemic was strongly associated with a poorer HRQoL (aOR=3.1; 95%CI: 2.3-4.4), especially psychosocial (aOR=3.2; 95%CI: 2.4-4.3) after adjustment for age, sex and health condition (Fig. 2). Children who experienced a severe pandemic impact were also more likely to present a poor mental health (aOR=3.9; 95%CI: 2.5-6.2), particularly internalizing problems (aOR=4.2; 95%CI: 2.6-6.6). Analyses performed with imputed data or with a different threshold for a severe pandemic impact retrieved comparable results (Additional file, table 4). When stratified by sex, having been severely impacted by the pandemic was more specifically associated with internalizing problems among girls and with externalizing problems among boys, although the interaction between the pandemic impact and sex was not significant (P-value>0.05, Additional file, table 5). The association between having been severely impacted by the pandemic and HRQoL or mental health remained significant, even after further adjustment for potential lifestyle or family confounders (Additional file, tables 6&7).

## DISCUSSION

In this study, 13% of children and adolescents had been severely impacted by the COVID-19 pandemic, which corresponds to a milder average impact than in previous reports (mean score: 3.7 vs 8.2-12.0) (17). Health and family vulnerabilities, as well as lifestyle changes because of the COVID-19 pandemic, resulted in a higher likelihood of experiencing a severe multidimensional impact. In turn, a severe impact of the pandemic was associated with a decreased HRQoL and poor mental health.

As previously observed, we found that children with a lasting physical or mental health condition were particularly affected during the pandemic (4,5). The consequences of healthcare disruptions related to the pandemic could have been direct on those children’s health and well-being, but also indirect through an increased family strain, since more care responsibilities fell on caregivers (6). Worries and self-imposed restrictions linked to a potential higher risk of SARS-CoV-2 infection among vulnerable children could have represented an additional burden.

In accordance with previous findings (25,26), unfavourable family circumstances were a determinant of experiencing a severe impact of the pandemic, with children living in households with disadvantaged socio-economic conditions, poor parent’s psychological state or poor parent-child relationship being more at risk. In fact, families facing such difficulties may have been particularly vulnerable to the changes induced by the pandemic. For example, individuals with low financial resources were more likely to lose their jobs (27) than those with a good financial situation. Likewise, adults with existing poor mental health seemed at risk of experiencing a worsening of their psychological state during the pandemic (28). In parallel, the pandemic context may have favoured such adverse conditions to spill over into the parent-child relationship. Indeed, sanitary measures heavily relied on and stressed the role of the household, with the disruption of childcare and healthcare, cancellations of extra-curricular activities and quarantines. Although such circumstances may have strengthened bonds within well-functioning families, they may have been challenging in families experiencing difficult situations (8). Overall, this reflects the concerns about the role of the COVID-19 pandemic in exacerbating social inequalities (29).

Recent research advocated that maintaining a daily routine, including a sufficient level of physical activity (30), was a good practice to mitigate the adverse effect of the pandemic. Our results echo these findings, since lifestyle and family changes such as a decrease or even an increase in time spent with parents was associated with a severe pandemic impact, as was a decrease in physical activity and an increase in screen time.

Interestingly, previous infection, evaluated with the anti-SARS-CoV-2 N serological status, was not associated with a severe pandemic impact, which concurs with observations from Kılınçel et al., who found no difference in HRQoL or anxiety among COVID-19-positive children, when compared with healthy controls. These results reflect the usually mild course of COVID-19 among young people (32) and suggest that, in general, the pandemic context had more impact on children’s HRQoL and mental health than the COVID-19 disease itself.

HRQoL level was higher in our results compared with international population-based studies conducted during the pandemic (mean PedsQL score: 83.8 vs 61.3-79.7) (33–35). Similarly, in our sample, occurrence of mental health problems was lower than reported in other countries (mean SDQ score: 7.2 vs 10.7-12.1) (35–38). Reasons could include higher level of HRQoL and lower prevalence of mental health problems observed in Swiss children already prior to the pandemic in comparison with other European countries (39). It may also be the result of milder consequences of the pandemic on youth’s well-being linked to the relatively lighter sanitary measures (40).

We showed that having been severely impacted by the pandemic was strongly associated with decreased HRQoL and mental health, which reflects findings from other population-based studies (38,41). The relationship was particularly pronounced with psychosocial HRQoL and internalizing problems, which highlights the impact of the pandemic on the psychological well-being of children, and echoes the increase in depression and anxiety observed worldwide (11). Moreover, it is important to consider that our findings relate to a country where relatively light restrictions were implemented throughout the pandemic, and where all measures have been eased two years after the first declared case. While most children seemed to fare well, the pandemic may have persistent indirect consequences in terms of quality of life and mental health even in Switzerland and with light restrictions, which is mirrored by the observed increase in demand for mental healthcare (42).

This study provides additional information for decision-makers to help balance infection control and young population’s well-being. Given the potential persistent consequences of pandemic-related stressors on the well-being of children, future measures should attempt to avoid heavy disruptions in their everyday life (43). If the epidemiological situation requires strong restrictions, support should be provided to mitigate the pandemic impact, especially for children with existing social or health vulnerabilities (44). It could include accessible and anonymous psychological counselling, the promotion of open communication and the practice of common activities within families, as well as the fostering of a healthy routine including physical activity (30,44). Since the family context also plays an essential role in children’s well-being, measures should aim in parallel at reducing the burden of the pandemic on parents. This could include flexible working time when possible, the availability of childcare services, counselling and support from healthcare workers, or financial aid for economically vulnerable families.

This study presents limitations. First, due to the cross-sectional design, the direction of the associations between socio-demographic or family characteristics, a severe impact of the pandemic and HRQol or mental health are only assumed but cannot be determined with certainty. We cannot exclude that reverse causation effects are at play. For example, having been severely impacted by the pandemic may have led to a worsening of the family and socio-economic situation; similarly, a poor pre-existing HRQoL or mental health could have been a determinant of a severe pandemic impact. However, the main conclusion of our study holds, that is the need for tailored support for vulnerable populations remains. Second, despite the random selection process, individuals with favourable socio-economic conditions were more likely to participate. This could have led to an underestimation of the prevalence of our outcomes since their occurrence was more common among underprivileged individuals. Third, one of the benefits of the Coronavirus impact scale lies in the inclusion of various domains at the individual and household level, which are translated into a multidimensional measure of impact. However, the perception of the severity of the impact of the pandemic may differ according to personal traits (45) and remains therefore highly subjective. Finally, we acknowledge that the pandemic may have positively impacted some children, although this was not examined due to the design of the coronavirus impact scale. Our study also has several strengths, the major being its reliance on a large sample of randomly selected children and adolescents covering a wide age range. It was also possible to examine multiple determinants of a severe impact of the pandemic at the individual and family level. Finally, validated HRQoL of life and mental health scales were used, enabling international comparisons.

## CONCLUSION

The COVID-19 pandemic may have persistent consequences on the quality of life of children and adolescents, especially among those with health and family vulnerabilities. Future measures disrupting their everyday life should be avoided and tailored support, including accessible mental healthcare, should be provided at the child and family level to promote their well-being.

## Supporting information

Strobe guidelines

## Data Availability

All data produced in the present study are available upon reasonable request to the authors

## DECLARATIONS

### Ethics approval and consent to participate

The Geneva Cantonal Commission for Research Ethics gave ethical approval for this work (IDs 2020-00881 & 2021-01973). Parents of participants and adolescents aged 14 or older provided written informed consent. Children gave oral assent to participate.

### Competing interests

The authors have no relevant financial or non-financial interests to disclose.

### Funding

The SEROCoV-KIDS study was funded by the Federal Office of Public Health of Switzerland and the Jacobs Foundation.

### Authors’ contributions

All authors contributed to the study conception and design. Material preparation and data collection were performed by Viviane Richard, Roxane Dumont, Elsa Lorthe, Andrea Loizeau, Hélène Baysson, Maria-Eugenia Zaballa and Francesco Pennacchio, under the supervision of Rémy Barbe, Klara Posfay-Barbe, Idris Guessous and Silvia Stringhini. Analyses were performed by Viviane Richard who also wrote the first draft of the manuscript. All authors critically revised the previous versions of the manuscript. All authors read and approved the final manuscript.

## Acknowledgments

We are grateful to the staff of the Unit of Population Epidemiology of the Division of Primary Care Medicine of the University Hospitals of Geneva, as well as to all participants whose contributions were invaluable to the study.

## Notes

### Competing Interest Statement

The authors have declared no competing interest.

### Author Declarations

The Geneva Cantonal Commission for Research Ethics gave ethical approval for this work (IDs 2020-00881 & 2021-01973).

