## Supplementary material for "Impact of the COVID-19 pandemic on children and adolescents: determinants and association with quality of life and mental health – A cross-sectional study"

Child and Adolescent Psychiatry and Mental Health - Supplementary material

Viviane Richard[a] M.Sc., Roxane Dumont[a] M.Sc., Elsa Lorthe[a] RM, Ph.D., Andrea Loizeau[a] Ph.D., Hélène Baysson[a,b], María-Eugenia Zaballa[a] Ph.D., Francesco Pennacchio[a] Ph.D. Ph.D., Rémy P. Barbe[c] M.D., Klara M. Posfay-Barbe[d] M.D., Ph.D., Idris Guessous[b,e]* M.D., Ph.D., Silvia Stringhini[a,b,f]* Ph.D.; for the SEROCoV-KIDS study group

* Authors contributed equally to this work.

### Affiliations

[a] Unit of Population Epidemiology, Division of Primary Care Medicine, Geneva University Hospitals, Jean-Violette 29, 1205 Geneva, Switzerland

[b] Department of Health and Community Medicine, Faculty of Medicine, University of Geneva, Michel-Servet 1, 1206 Geneva, Switzerland

[c] Division of Child and Adolescent Psychiatry, Department of Woman, Child, and Adolescent Medicine, Geneva University Hospitals, Willy Donzé 6, 1211 Geneva, Switzerland

[d] Division of General Pediatrics, Department of Woman, Child, and Adolescent Medicine, Geneva University Hospitals, Willy Donzé 6, 1211 Geneva, Switzerland

[e] Division and Department of Primary Care Medicine, Geneva University Hospitals, Gabrielle-Perret-Gentil 4, 1205 Geneva, Switzerland

[f] University Center for General Medicine and Public Health, University of Lausanne, Bugnon 44, 1011 Lausanne, Switzerland

### SEROCoV-KIDS study group

Deborah Amrein, Isabelle Arm-Vernez, Andrew S Azman, Antoine Bal, Michael Balavoine, Rémy P Barbe, Hélène Baysson, Julie Berthelot, Patrick Bleich, Livia Boehm, Gaëlle Bryand, Viola Bucolli, Prune Collombet, Alain Cudet, Vladimir Davidovic, Carlos de Mestral Vargas, Paola D'Ippolito, Richard Dubos, Roxane Dumont, Isabella Eckerle, Nacira El Merjani, Marion Favier, Natalie Francioli, Clément Graindorge, Idris Guessous, Séverine Harnal, Samia Hurst, Laurent Kaiser, Omar Kherad, Julien Lamour, Pierre Lescuyer, Arnaud G. L'Huillier, Andrea Jutta Loizeau, Elsa Lorthe, Chantal Martinez, Stéphanie Mermet, Mayssam Nehme, Natacha Noël, Francesco Pennacchio, Javier Perez-Saez, Anne Perrin, Didier Pittet, Klara M Posfay-Barbe, Jane Portier, Géraldine Poulain, Caroline Pugin, Nick Pullen, Viviane Richard, Frederic Rinaldi, Deborah Rochat, Cyril Sahyoun, Irine Sakvarelidze, Khadija Samir, Hugo Alejandro Santa Ramirez, Jessica Rizzo, Stephanie Schrempft, Claire Semaani, Silvia Stringhini, Stéphanie Testini, Yvain Tisserand, Deborah Urrutia Rivas, Charlotte Verolet, Jennifer Villers, Guillemette Violot, Nicolas Vuilleumier, Sabine Yerly, María-Eugenia Zaballa, Christina Zavlanou

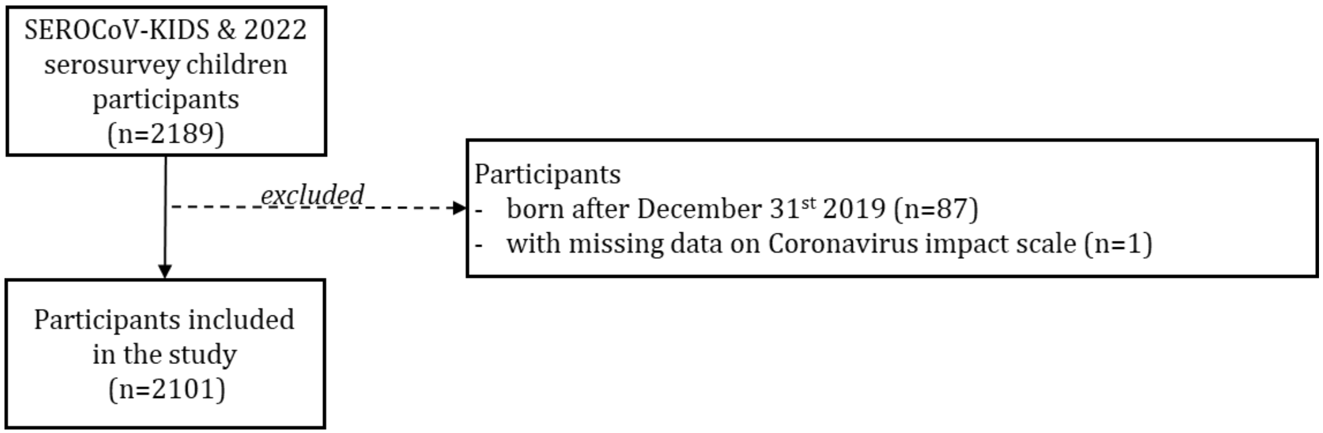


Figure 1. Flow chart of the study participants

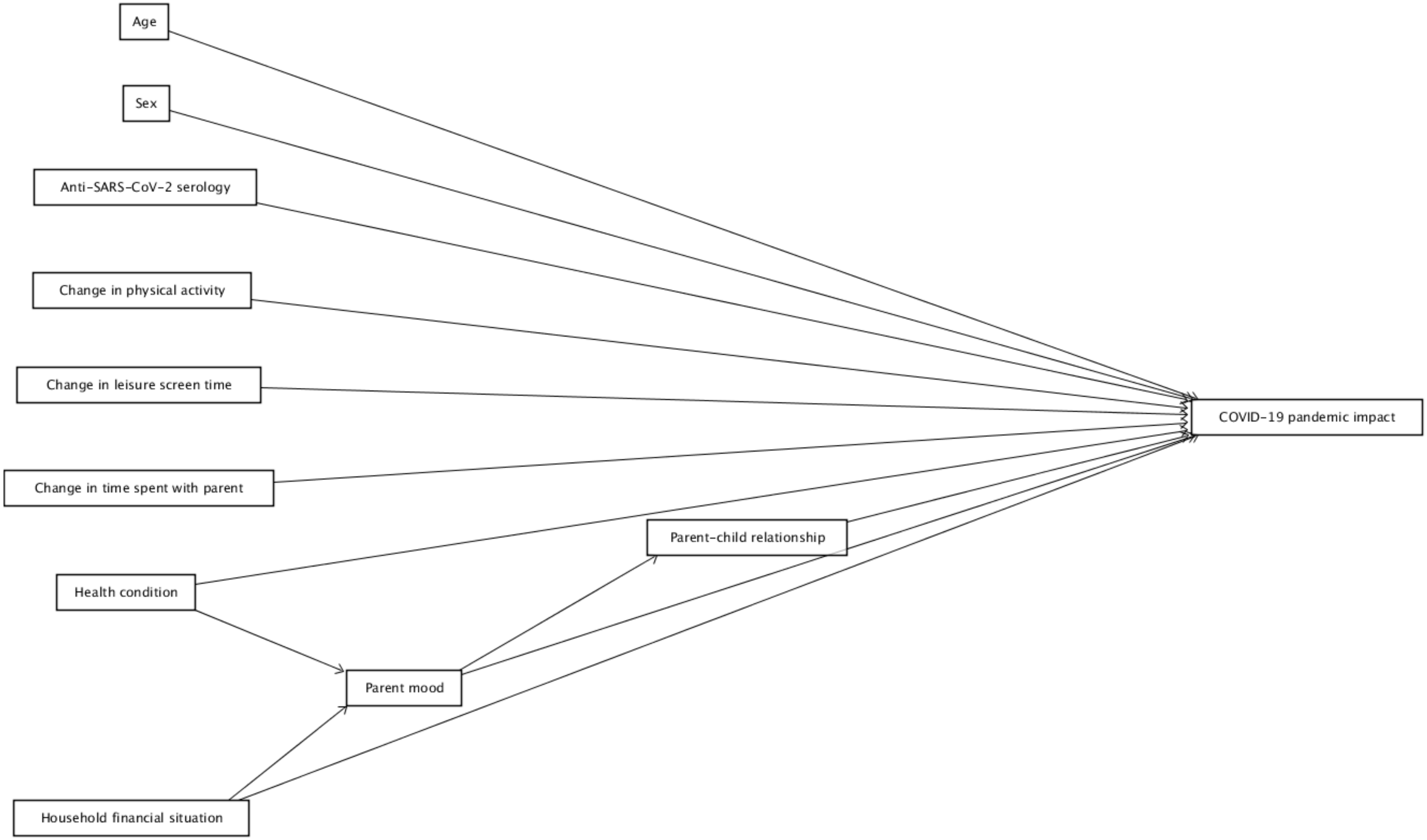


Figure 2. Directed acyclic graph of the potential determinants of the COVID-19 pandemic impact. Age and sex were deemed covariates for all other variables (see Methods); arrows are not drawn for readability purpose.

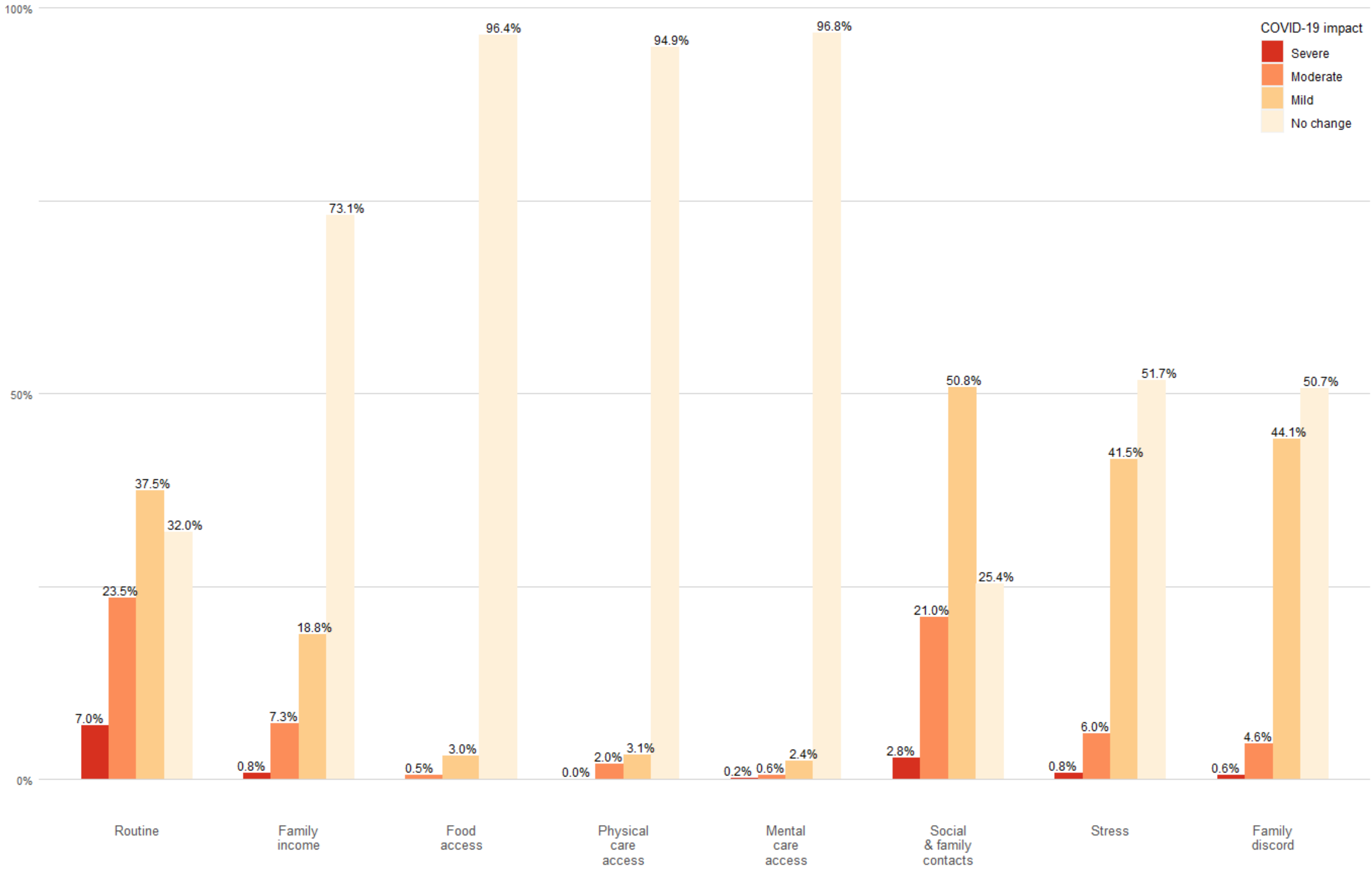


Figure 3. Individual item responses to the Coronavirus Impact scale. Parents reported on behalf of their children how each domain had been impacted by the COVID-19 pandemic, n=2101.

Table 1. Physical and mental health condition distribution in the sample as reported by parents (n=2101)

| **Physical disease or condition** | **n** | **%** | **Mental behavioural and neurodevelopmental disorder** | **n** | **%** |
|---|---|---|---|---|---|
| No physical disease or condition | 1797 | 85.5 | No mental disease or disorder | 1913 | 91.1 |
| Chromosomal abnormality | 4 | 0.2 | Developmental delay | 8 | 0.4 |
| Asthma | 84 | 4.0 | Anxiety disorder | 24 | 1.1 |
| Other lung disease | 9 | 0.4 | Learning disability | 83 | 4.0 |
| Diabetes | 2 | 0.1 | Attention deficit hyperactivity disorder | 75 | 3.6 |
| Other endocrine disease | 8 | 0.4 | Autism spectrum disorder | 15 | 0.7 |
| Epilepsy | 4 | 0.2 | Eating disorder | 11 | 0.5 |
| Heavy or frequent headache, migraine | 42 | 2.0 | Mood disorder | 9 | 0.4 |
| Physical disability | 6 | 0.3 | Gender identity disorder | 1 | 0.0 |
| Haematological disease | 8 | 0.4 | Obsessive-compulsive disorder | 2 | 0.1 |
| Rheumatic disease | 4 | 0.2 | Other mental disease or disorder | 19 | 0.9 |
| Disease or malformation of the skeleton, joints, or muscles | 24 | 1.1 | | | |
| Inflammatory bowel disease | 4 | 0.2 | | | |
| Other bowel disease | 6 | 0.3 | | | |
| Gastroesophageal reflux | 11 | 0.5 | | | |
| Overweight or obesity | 20 | 1.0 | | | |
| Hearing problem | 15 | 0.7 | | | |
| Heart disease or malformation | 13 | 0.6 | | | |
| Renal or genitourinary disease or malformation | 13 | 0.6 | | | |
| Immune disorder | 5 | 0.2 | | | |
| Other physical disease or condition | 66 | 3.1 | | | |

Results are numbers (n) and percentages (%) of children and adolescents with physical or mental health condition. Parents could report more than one condition and thus percentages do not add up to 100%.

Table 2. Outcomes comparison according to recruitment period (n=2101)

| | Before measures lifting | After measures lifting | |
|---|---|---|---|
| | **Median (IQR)** | **Median (IQR)** | **P-value** |
| Coronavirus impact score | 3.0 (2.0 - 5.0) | 3.0 (2.0 - 5.0) | 1.000 |
| Health-related quality of life (PedsQL total score) | 86.7 (77.1 - 95.0) | 86.7 (75.0 - 93.3) | 0.057 |
| Mental health (SDQ total score) | 6.0 (3.0 - 10.0) | 6.0 (4.0 - 10.0) | 0.021 |

Results are median (interval interquartile) and p-values are from Kruskal-Wallis test adjusted for multiple comparison with the Bonferroni method. COVID-19-related sanitary measures were lifted on February 3rd 2022 in Switzerland.

Table 3. Determinants of a severe COVID-19 pandemic impact score among children and adolescents. Sensitivity analyses with data imputation and severe impact definition.

| | | Severe pandemic impact defined as > mean + SD | | Severe pandemic impact defined as highest tertile | |
|---|---|---|---|---|---|
| | | Complete case analyses (n=2043) | Analyses with imputed data [a] (n=2098) | Complete case analyses (n=2043) | Analyses with imputed data [a] (n=2098) |
| | Level | aOR (95% CI) | aOR (95% CI) | aOR (95% CI) | aOR (95% CI) |
| Age | Years | 1.03 (0.99 - 1.06) | - | 1.03 (1.00 - 1.06) | - |
| Sex [b] (ref. female) | Male | 0.92 (0.71 - 1.19) | - | 0.94 (0.76 - 1.15) | - |
| Health condition (ref. no) | Yes | 1.85 (1.38 - 2.48) | 1.88 (1.41 - 2.50) | 1.92 (1.51 - 2.45) | 1.96 (1.55 - 2.48) |
| Anti-SARS-CoV-2 serological status [c] (ref. negative) | Positive | 0.91 (0.69 - 1.22) | 0.90 (0.68 - 1.20) | 1.05 (0.83 - 1.33) | 1.01 (0.80 - 1.27) |
| | Undetermined | 0.42 (0.06 - 3.01) | 0.39 (0.05 - 2.96) | 0.83 (0.25 - 2.75) | 0.75 (0.22 - 2.51) |
| Friends (ref. several) | One | 1.87 (1.33 - 2.62) | - | 1.55 (1.17 - 2.06) | - |
| | None | 3.22 (1.76 - 5.90) | - | 2.39 (1.37 - 4.15) | - |
| | Unknown / na | 1.49 (0.89 - 2.48) | - | 1.40 (0.95 - 2.08) | - |
| Change in physical activity [d,e] (ref. similar or increase) | Decrease | 3.86 (2.85 - 5.23) | - | 4.11 (3.17 - 5.34) | - |
| | Unknown / na | 1.61 (1.03 - 2.50) | - | 1.74 (1.23 - 2.45) | - |
| Change in leisure screen time [d,e] (ref. similar or decrease) | Increase | 1.98 (1.48 - 2.64) | 1.98 (1.48 - 2.64) | 2.27 (1.79 - 2.87) | 2.29 (1.81 - 2.89) |
| | Unknown / na | 0.78 (0.42 - 1.47) | 0.87 (0.48 - 1.57) | 1.14 (0.73 - 1.78) | 1.21 (0.78 - 1.86) |
| Change in time spent with parent [d] (ref. similar) | Increase | 2.08 (1.54 - 2.80) | - | 2.13 (1.69 - 2.69) | - |
| | Decrease | 6.48 (4.08 - 10.28) | - | 5.35 (3.45 - 8.31) | - |
| | Unknown | 6.58 (3.57 - 12.10) | - | 5.04 (2.83 - 8.97) | - |
| Parent-child relationship (ref. good) | Rather good | 1.85 (1.34 - 2.57) | 1.89 (1.38 - 2.59) | 1.79 (1.38 - 2.34) | 1.82 (1.40 - 2.36) |
| | Average-to-poor | 4.64 (2.26 - 9.52) | 4.24 (2.10 - 8.53) | 5.66 (2.78 - 11.53) | 5.56 (2.79 - 11.07) |
| Parent mood (ref. good) | Average-to-poor | 2.98 (2.16 - 4.10) | 2.86 (2.09 - 3.93) | 2.38 (1.79 - 3.18) | 2.28 (1.72 - 3.03) |
| Household financial situation (ref. very good) | Good | 1.76 (1.23 - 2.50) | 1.79 (1.27 - 2.54) | 1.69 (1.29 - 2.21) | 1.74 (1.33 - 2.26) |
| | Average-to-poor | 4.63 (3.18 - 6.75) | 4.73 (3.26 - 6.87) | 3.22 (2.37 - 4.39) | 3.28 (2.42 - 4.45) |
| | No answer | 1.24 (0.64 - 2.39) | 1.30 (0.69 - 2.46) | 1.15 (0.69 - 1.90) | 1.17 (0.71 - 1.91) |

Results are adjusted odds ratios (aOR) with 95% confidence intervals (95% CI) from generalized estimating equations taking the household clustering into account, adjusted for age and sex. Model for parent mood adjusted for age, sex, health condition and financial situation; model for parent-child relationship adjusted for age, sex and parent mood. Not applicable (na) if stated so by parents or if the child was younger than 6 years at questionnaire completion for friends, than 1 year at pandemic onset for leisure screen time change or than 2 years at pandemic onset for physical activity change.
[a] Missing data imputed using chained equations, see methods. [b] The sex category "Other" was not included because of too small number of observations. [c] Complete case analysis based on participants with available anti-SARS-CoV-2 serology (N=1966). [d] participant-reported change as a result of the COVID-19 pandemic. [e] Similar level and increase in physical activity as well as similar level and decrease in screen time are grouped. When analyzed separately; coefficients were of comparable magnitude.

Table 4. Association between having been severely impacted by the pandemic and health-related quality of life (HRQoL) or mental health of children and adolescents. Sensitivity analyses with data imputation and severe impact definition.

| | Severe pandemic impact defined as > mean + SD | | Severe pandemic impact defined as highest tertile | |
|---|---|---|---|---|
| | **Complete case analyses (n=2043)** | **Analyses with imputed data (n=2098)** | **Complete case analyses (n=2043)** | **Analyses with imputed data (n=2098)** |
| **Severe pandemic impact score on:** | **aOR (95% CI)** | **aOR (95% CI)** | **aOR (95% CI)** | **aOR (95% CI)** |
| Poor overall HRQoL | 3.14 (2.26 - 4.35) | 3.12 (2.25 - 4.32) | 2.44 (1.82 - 3.27) | 2.43 (1.82 - 3.26) |
| Poor psychosocial HRQoL | 3.20 (2.40 - 4.25) | 3.07 (2.30 - 4.09) | 2.65 (2.07 - 3.40) | 2.57 (2.00 - 3.29) |
| Poor physical HRQoL | 2.06 (1.33 - 3.20) | 2.14 (1.38 - 3.31) | 1.59 (1.08 - 2.32) | 1.63 (1.11 - 2.38) |
| Poor overall mental health | 3.93 (2.49 - 6.20) | 3.72 (2.38 - 5.83) | 3.56 (2.32 - 5.46) | 3.62 (2.38 - 5.51) |
| Internalizing problems | 4.16 (2.64 - 6.56) | 4.15 (2.63 - 6.55) | 3.85 (2.47 - 6.00) | 3.98 (2.59 - 6.13) |
| Externalizing problems | 2.36 (1.40 - 3.97) | 2.38 (1.44 - 3.94) | 2.62 (1.67 - 4.13) | 2.66 (1.71 - 4.14) |

Results are adjusted odds ratios (aOR) with 95% confidence intervals (95% CI) from generalized estimating equations taking the household clustering into account and adjusted for age, sex and health condition. Missing data on HRQoL and mental health imputed using chained equations (see methods).

Table 5. Association between having been severely impacted by the pandemic and health-related quality of life (HRQoL) or mental health of children and adolescents, stratified by sex

| | **Girls (n=1013)** | **Boys (n=1030)** |
|---|---|---|
| **Severe pandemic impact score on:** | **aOR (95% CI)** | **aOR (95% CI)** |
| Poor overall HRQoL | 3.56 (2.33 - 5.43) | 2.81 (1.71 - 4.61) |
| Poor psychosocial HRQoL | 3.12 (2.10 - 4.65) | 3.29 (2.19 - 4.92) |
| Poor physical HRQoL | 1.95 (1.05 - 3.63) | 2.00 (1.00 - 3.99) |
| Poor overall mental health | 3.96 (1.97 - 7.96) | 3.95 (2.15 - 7.26) |
| Internalizing problems | 4.58 (2.55 - 8.25) | 2.98 (1.14 - 7.78) |
| Externalizing problems | 1.24 (0.41 - 3.70) | 3.14 (1.70 - 5.80) |

Results are adjusted odds ratios (aOR) with 95% confidence intervals (95% CI) from generalized estimating equations taking the household clustering into account and adjusted for age, sex and health condition. Severe pandemic impact defined as a score > mean + 1SD.

Table 6. Association between the COVID-19 pandemic impact score and the health-related quality of life of children of adolescents according to different adjustment strategies (n=2043)

| | | Demographic | Demographic + Health | Demographic + Health + Lifestyle | Demographic + Health + Family | Demographic + Health + Lifestyle + Family |
|---|---|---|---|---|---|---|
| | | aOR (95% CI) | aOR (95% CI) | aOR (95% CI) | aOR (95% CI) | aOR (95% CI) |
| **Severe pandemic impact (ref. none or mild)** | **Severe** | **3.32 (2.40 - 4.59)** | **3.14 (2.26 - 4.35)** | **2.64 (1.89 - 3.69)** | **2.00 (1.40 - 2.85)** | **1.80 (1.25 - 2.58)** |
| Age | Years | 1.13 (1.09 - 1.17) | 1.12 (1.08 - 1.16) | 1.12 (1.06 - 1.18) | 1.11 (1.07 - 1.16) | 1.12 (1.06 - 1.18) |
| Sex [a] (ref. female) | Male | 0.76 (0.58 - 1.00) | 0.76 (0.57 - 1.00) | 0.75 (0.57 - 1.00) | 0.76 (0.57 - 1.01) | 0.75 (0.56 - 1.01) |
| Health condition (ref. no) | Yes | | 1.70 (1.25 - 2.30) | 1.61 (1.18 - 2.20) | 1.55 (1.13 - 2.14) | 1.51 (1.09 - 2.08) |
| Friends (ref. several) | One | | | 1.57 (1.10 - 2.23) | | 1.39 (0.96 - 2.01) |
| | None | | | 2.43 (1.22 - 4.85) | | 2.05 (1.02 - 4.09) |
| | Unknown / na | | | 1.49 (0.77 - 2.87) | | 1.50 (0.77 - 2.93) |
| Change in physical activity time (ref. similar or increase) | Decrease | | | 1.23 (0.86 - 1.74) | | 1.13 (0.78 - 1.64) |
| | Unknown / na | | | 0.92 (0.52 - 1.62) | | 0.84 (0.47 - 1.51) |
| Change in leisure screen time (ref. similar or decrease) | Increase | | | 1.43 (1.05 - 1.97) | | 1.41 (1.01 - 1.95) |
| | Unknown / na | | | 1.15 (0.61 - 2.18) | | 1.16 (0.60 - 2.24) |
| Change in time spent with parent (ref. similar) | Increase | | | | 1.05 (0.75 - 1.46) | 0.99 (0.70 - 1.40) |
| | Decrease | | | | 2.10 (1.22 - 3.62) | 1.85 (1.09 - 3.15) |
| | Unknown | | | | 1.35 (0.65 - 2.82) | 1.27 (0.60 - 2.68) |
| Parent-child relationship (ref. good) | Rather good | | | | 1.88 (1.33 - 2.66) | 1.81 (1.28 - 2.58) |
| | Average-to-poor | | | | 2.78 (1.48 - 5.25) | 2.78 (1.45 - 5.34) |
| Parent mood (ref. good) | Average-to-poor | | | | 1.66 (1.11 - 2.47) | 1.67 (1.12 - 2.50) |
| Household financial situation (ref. very good) | Good | | | | 1.70 (1.17 - 2.47) | 1.67 (1.15 - 2.43) |
| | Average-to-poor | | | | 2.29 (1.50 - 3.50) | 2.20 (1.43 - 3.38) |
| | No answer | | | | 1.72 (0.89 - 3.29) | 1.75 (0.93 - 3.31) |

Results are adjusted odds ratios (aOR) with 95% confidence intervals (95% CI) from generalized estimating equations taking the household clustering into account. Severe pandemic impact defined as a score > mean + 1SD. Not applicable (na) if stated so by parents or if the child was younger than 6 years at questionnaire completion for friends, than 1 year at pandemic onset for leisure screen time change or than 2 years at pandemic onset for physical activity change. [a] The sex category "Other" was not included because of too small number of observations.

Table 7. Association between the COVID-19 pandemic impact score and the mental health of children of adolescents according to different adjustment strategies (n=2043)

| | | Demographic | Demographic + Health | Demographic + Health + Lifestyle | Demographic + Health + Family | Demographic + Health + Lifestyle + Family |
|---|---|---|---|---|---|---|
| | | **aOR (95% CI)** | **aOR (95% CI)** | **aOR (95% CI)** | **aOR (95% CI)** | **aOR (95% CI)** |
| **Severe pandemic impact (ref. mild)** | **Severe** | **4.53 (2.94 - 6.97)** | **3.93 (2.49 - 6.20)** | **3.01 (1.86 - 4.87)** | **2.85 (1.68 - 4.85)** | **2.45 (1.40 -4.30)** |
| Age | Years | 0.97 (0.92 - 1.02) | 0.93 (0.88 - 0.98) | 0.95 (0.88 - 1.04) | 0.88 (0.84 - 0.93) | 0.91 (0.84 - 0.99) |
| Sex [a] (ref. female) | Male | 1.40 (0.93 - 2.12) | 1.36 (0.89 - 2.08) | 1.40 (0.91 - 2.16) | 1.47 (0.94 - 2.30) | 1.48 (0.94 - 2.34) |
| Health condition (ref. no) | Yes | | 4.73 (3.13 - 7.15) | 4.42 (2.89 - 6.77) | 5.44 (3.52 - 8.41) | 5.36 (3.44 - 8.36) |
| Friends (ref. several) | One | | | 3.56 (2.13 - 5.93) | | 3.56 (2.09 - 6.07) |
| | None | | | 5.00 (2.06 - 12.16) | | 4.59 (1.58 - 13.34) |
| | Unknown / na | | | 2.47 (1.02 - 5.98) | | 2.24 (0.91 - 5.54) |
| Change in physical activity time (ref. similar or increase) | Decrease | | | 1.48 (0.86 - 2.55) | | 1.37 (0.73 - 2.57) |
| | Unknown / na | | | 1.56 (0.77 - 3.19) | | 1.60 (0.79 - 3.28) |
| Change in leisure screen time (ref. similar or decrease) | Increase | | | 1.11 (0.69 - 1.80) | | 1.04 (0.64 - 1.70) |
| | Unknown / na | | | 0.65 (0.25 - 1.67) | | 0.66 (0.26 - 1.63) |
| Change in time spent with parent (ref. similar) | Increase | | | | 1.57 (0.96 - 2.55) | 1.56 (0.96 - 2.55) |
| | Decrease | | | | 1.77 (0.91 - 3.41) | 1.42 (0.71 - 2.82) |
| | Unknown | | | | 1.33 (0.38 - 4.64) | 1.02 (0.27 - 3.90) |
| Parent-child relationship (ref. good) | Rather good | | | | 5.36 (3.30 - 8.70) | 5.36 (3.24 - 8.87) |
| | Average-to-poor | | | | 14.28 (6.27 - 32.49) | 14.05 (5.72 - 34.49) |
| Parent mood (ref. good) | Average-to-poor | | | | 0.94 (0.52 - 1.69) | 1.00 (0.54 - 1.83) |
| Household financial situation (ref. very good) | Good | | | | 1.38 (0.81 - 2.33) | 1.27 (0.75 - 2.16) |
| | Average-to-poor | | | | 1.09 (0.54 - 2.18) | 0.85 (0.41 - 1.77) |
| | No answer | | | | 1.47 (0.53 - 4.10) | 1.66 (0.61 - 4.50) |

Results are adjusted odds ratios (aOR) with 95% confidence intervals (95% CI) from generalized estimating equations taking the household clustering into account. Severe pandemic impact defined as a score > mean + 1SD. Not applicable (na) if stated so by parents or if the child was younger than 6 years at questionnaire completion for friends, than 1 year at pandemic onset for leisure screen time change or than 2 years at pandemic onset for physical activity change. [a] The sex category "Other" was not included because of too small number of observations.
